# Healthcare Cost Literacy: Exploration of Concept and Initial Development of a Novel Tool in a Representative U.S. Adult Population

**DOI:** 10.1101/2023.12.08.23292892

**Authors:** Alexandra Guttentag, Haider J Warraich, Jeroen van Meijgaard

## Abstract

There is a growing need for understanding and addressing the issue of healthcare cost literacy (HCL) in the United States. We conducted a survey in partnership with YouGov targeting a representative sample of 1500 American adults (median age 47, 51.4% female, 62.8% white) to help develop a novel tool to assess the prevalence of HCL, and to estimate levels of HCL and health literacy across various sociodemographic and health-related variables. An exploratory factor analysis revealed that the HCL questions mapped to three factors: 1) knowledge on health insurance terminology/interpretation, 2) ability to estimate healthcare costs ahead of time, and 3) confidence in performing cost comparisons between healthcare plans and deductibles. An understanding of Americans’ levels of HCL will help policymakers and various stakeholders in the healthcare system to develop targeted plans to educate consumers on financial planning and evaluation in the healthcare system.

## Background

Health literacy (HL) has long been recognized as a critical determinant of health outcomes and healthcare utilization and decision-making across different population groups^1–3^. HL is defined by the Centers for Disease Control and Prevention (CDC) as “the ability to find, understand, and use information and services to inform health-related decisions and actions for themselves and others”^4,5^. And over the last few decades, researchers and practitioners have increasingly focused on strategies to enhance HL among individuals and communities^6–8^.

The escalating costs of healthcare have recently emerged as a major challenge resulting in individuals and families being unable to bear the financial burden of medical expenses^9–11^. Consequently, there is a growing need for understanding and addressing the issue of healthcare *cost* literacy (HCL). While there is emergent data estimating the prevalence of health literacy and financial burden from healthcare, little is known about the prevalence of and factors determining HCL and its relationship with conventionally assessed HL. Therefore, we conducted a comprehensive survey targeting a representative sample of American adults to help devise a novel definition of HCL, to develop a tool to help assess the prevalence of HCL, and to estimate the relationship between HL and HCL across various sociodemographic and health-related variables. The goal of this research is to present an overview of the initial development of the HCL tool and discuss how it might be applied in different research settings to understand literacy around the costs of healthcare.

## Methods

The data for this study was collected in partnership with YouGov (www.YouGov.com), an online survey platform tool which comprises a proprietary panel of opt-in participants. The sample of participants for YouGov studies are recruited through different methods, including online advertising and email invitations. Panelists have previously given consent to be contacted periodically for participation in surveys. Incentives are provided for participants in the form of YouGov points, which can be redeemed for gift cards and other rewards. The sample for this study is nationally representative and weighted on age, gender, race/ethnicity, political affiliation, educational attainment, and region. The weights are generated using raking (or “iterative proportional fitting”) to match the target demographics, in this case the U.S. adult population.

We measured participants’ HL using the validated 12-point HLSQ-12 scale initially developed by Finbråten et al^12^. The HLS-Q12 is a short version of the 47-item European Health Literacy Survey Questionnaire and encompasses both cognitive domains (ability to access, understand, appraise, and apply health information) and health domains (knowledge of health care, disease prevention, and health promotion). Questions are asked on a scale of 1 (“very difficult”) to 4 (“very easy”); a higher score indicates higher HL proficiencies. The total number of points on the HLSQ-12 could possibly range from 12 (answering “very difficult” for all of the questions) to 48 points (answering “very easy” to all 12 questions)^12^.

We adapted the CDC definition of HL and defined HCL as “the ability to find, understand and use information and services related to healthcare costs, enabling individuals to make informed decisions about their healthcare expenditure.” To estimate levels of HCL, we asked participants a series of questions related to their understanding of various costs associated with medical care – from knowledge about cost-differences in generic versus brand drugs to knowledge about hospital chargemasters and bill contesting, to confidence in being able to estimate healthcare bills prior to receiving services. We asked 12 HCL questions on a scale of 1 (“very difficult”) to 4 (“very easy”), which aligns with the scale provided in the HLSQ-12 instrument. Participants were asked a range of questions around HCL that spanned several conceptual domains including knowledge about costs of healthcare procedures, visits, and medications, and knowledge about various components of health insurance. Three additional questions related to HCL presented binary yes or no answer choices (“Did you know that different pharmacies may charge different prices for the same drug?”, “Did you know that the hospital chargemaster exists and can be requested/reviewed?”, and “Do you know how to contest a hospital bill?”).

HCL was analyzed in several different ways: as individual questions, as a continuous measure (by taking the average score on all 12 questions scored on 1-4 point Likert scale), and with a binary categorization (median split using summed score of HCL questions with 1-4 point Likert scale).

Missing data in the HCL Likert scale questions (12 total) and HLSQ-12 were partially imputed. If a participant responded to 75% or more of the questions in the HLSQ-12 or in the series of 12 HCL questions, we imputed any missing questions using the mean score of the answered questions. For participants who missed more than 25% of the questions, their scores on HL and/or HCL were coded as “Not Applicable” (N=89, or 6.0%, dropped for HLSQ-12, N=202, or 13.5%, dropped for HCL). HL and HCL were evaluated as covariates subsequent analyses using an individual’s mean response score across all 12 questions in each scale; thus, each respondent with 75% response rate for the HLSQ-12 has a mean HL score, and each respondent with at least 75% response rate across the HCL has a mean HCL score.

Participants also completed a five-item “assessment” of health insurance terms, which is adapted from survey(s) administered by the Understanding America Study, which is maintained by the Center for Economic and Social Research (CESR) at the University of Southern California^13^. We performed a bivariate analysis of the sum of correct answers on this assessment (scoring “Don’t know/NA” and incorrect responses as 0 and correct responses as 1 alongside other participant characteristics.

We performed an exploratory factor analysis (EFA) on the HL and HCL survey items to evaluate potential theoretical constructs underpinning the questions. EFA is employed in survey construction work to assess the underlying constructs (“factors”) of a dataset; these construct determinations are determined using inter-variable correlation metrics^14^. Typically, when an EFA is run on a dataset, a certain number of constructs are extracted and each variable is assigned to one of the constructs; from there, a researcher can qualitatively evaluate how each construct ties a certain group of variables together. For the EFA, only participants with full data were included; any participant missing any one variable included in the EFA was excluded. Sample sizes for complete cases were N=1,029 (HL), N=967 (HCL), and N=787 (HL and HCL combined). We performed separate EFAs on the HLSQ-12, the 12 HCL items, and the 24 HL and HCL items together. For all EFAs, we implemented a parallel analysis approach^14^, followed by EFA using oblique rotation and ordinary least squares specifying varying number of factors. All analyses were performed using R Version 4.3.1. The Advarra IRB determined that this research project was exempt from IRB oversight.

## Results

A total of 1500 complete survey responses were collected (median age 47, 51.4% female, 62.8% white). Sample weights were provided by the vendor (YouGov) using a sampling frame based upon the 2019 American Community Survey (ACS) data, and matching to age, gender, race/ethnicity, political affiliation, educational attainment, and region. Population characteristics (raw and weighted) of the sample are presented in **Table 1**.

**Table 1:** Weighted and unweighted characteristics of sample.

| **Table 1: Weighted and unweighted characteristics of sample** |  |  |
| --- | --- | --- |
| **Variable** | **Unweighted N (%)**<br>**(n=1,500)** | **Weighted %** |
| **Age** |  |  |
| 18-39 | 543 (36.2) | 38.1 |
| 40-64 | 634 (42.3) | 40.7 |
| 65+ | 323 (21.5) | 21.2 |
| **Gender** |  |  |
| Male | 695 (46.3) | 48.6 |
| Female | 805 (53.7) | 51.4 |
| **Race/Ethnicity** |  |  |
| Black | 189 (12.6) | 12.5 |
| Hispanic | 210 (14.0) | 16.0 |
| white | 928 (64.6) | 62.8 |
| Other | 132 (8.8) | 8.7 |
| **Educational Attainment** |  |  |
| High school or below | 558 (37.2) | 38.0 |
| Above high school | 942 (62.8) | 62.0 |
| **Employment Status** |  |  |
| Part Time Employed | 209 (14.0) | 13.5 |
| Full Time Employed | 573 (38.4) | 39.6 |
| Retired | 320 (21.4) | 21.0 |
| Unemployed | 107 (7.2) | 7.1 |
| Other | 283 (19.0) | 18.8 |
| **Household Income** |  |  |
| <30k | 313 (23.4) | 23.1 |
| 30-100k | 698 (52.1) | 52.9 |
| Over-100k | 328 (24.5) | 24.1 |
| **Insurance Status** |  |  |
| Employer-sponsored | 506 (33.8) | 35.1 |
| Exchange | 118 (7.9) | 7.6 |
| Medicaid | 223 (14.9) | 14.5 |
| Medicare | 338 (22.5) | 21.8 |
| Uninsured | 191 (12.7) | 13.1 |
| Other | 123 (8.2) | 8.0 |
| **Self-Rated Health Evaluation** |  |  |
| Excellent-Good | 1135 (73.1) | 76.3 |
| Fair-Poor | 358 (26.9) | 23.7 |

### HCL Tool Development

In the EFA on the 12-item theorized HCL questions, the Kaiser-Meyer-Olkin (KMO) factor adequacy provided an MSA of 0.94, and Bartlett’s test of sphericity provided a p-value <.001, supporting the EFA approach^15^. The parallel analysis suggested a three-factor model for the HLSQ-12, and a four-factor model for the HCL questions (**Tables 2a** and **2b**, respectively); when the HLSQ-12 and 12-item HCL questions were assessed together **(Table 2c)**, five factors were suggested. There was clear separation in factor assignment between the HL and HCL items, apart from one of the HL questions (Q7) which gets grouped with the HCL knowledge questions in the three-factor and four-factor cases.

**Table 2a.**
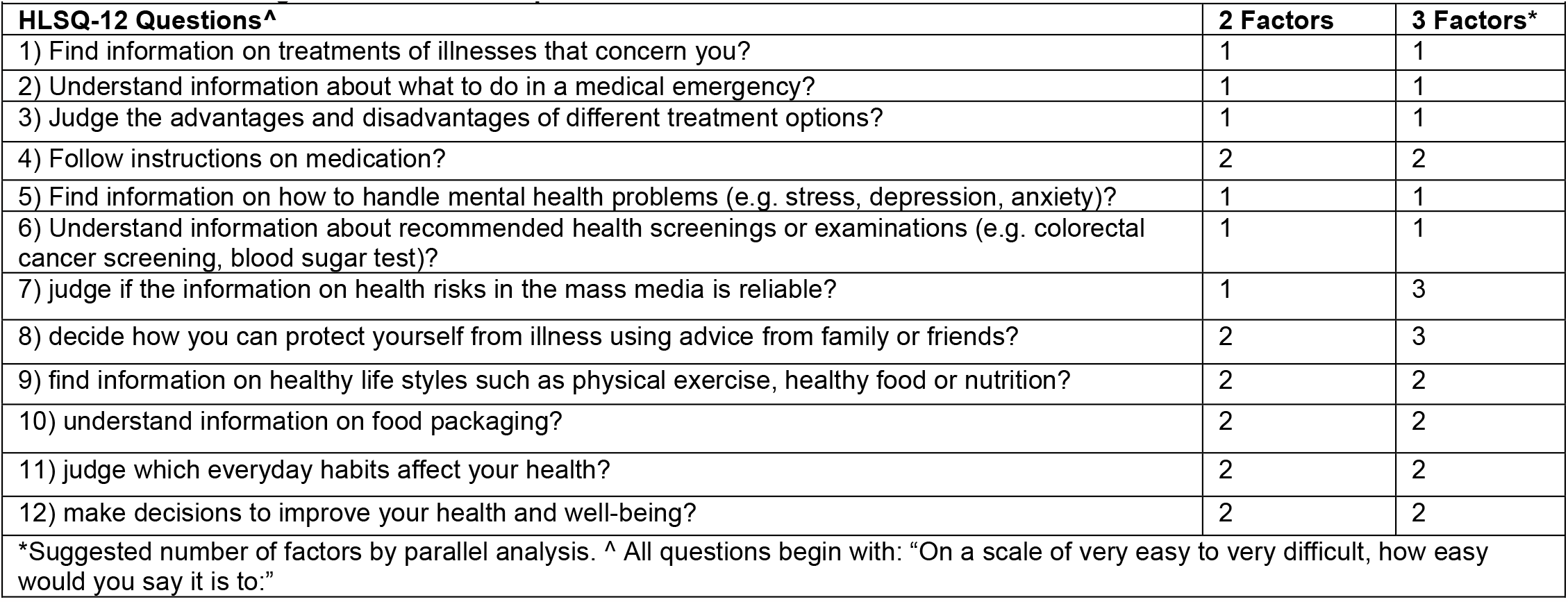
Factor assignment of HLSQ-12 questions.

| **Table 2a. Factor assignment of HLSQ-12 questions** |  |  |
| --- | --- | --- |
| **HLSQ-12 Questions^** | **2 Factors** | **3 Factors\*** |
| 1) Find information on treatments of illnesses that concern you? | 1 | 1 |
| 2) Understand information about what to do in a medical emergency? | 1 | 1 |
| 3) Judge the advantages and disadvantages of different treatment options? | 1 | 1 |
| 4) Follow instructions on medication? | 2 | 2 |
| 5) Find information on how to handle mental health problems (e.g. stress, depression, anxiety)? | 1 | 1 |
| 6) Understand information about recommended health screenings or examinations (e.g. colorectal cancer screening, blood sugar test)? | 1 | 1 |
| 7) judge if the information on health risks in the mass media is reliable? | 1 | 3 |
| 8) decide how you can protect yourself from illness using advice from family or friends? | 2 | 3 |
| 9) find information on healthy life styles such as physical exercise, healthy food or nutrition? | 2 | 2 |
| 10) understand information on food packaging? | 2 | 2 |
| 11) judge which everyday habits affect your health? | 2 | 2 |
| 12) make decisions to improve your health and well-being? | 2 | 2 |
| *Suggested number of factors by parallel analysis. ^ All questions begin with: "On a scale of very easy to very difficult, how easy would you say it is to:" |  |  |

**Table 2b.**
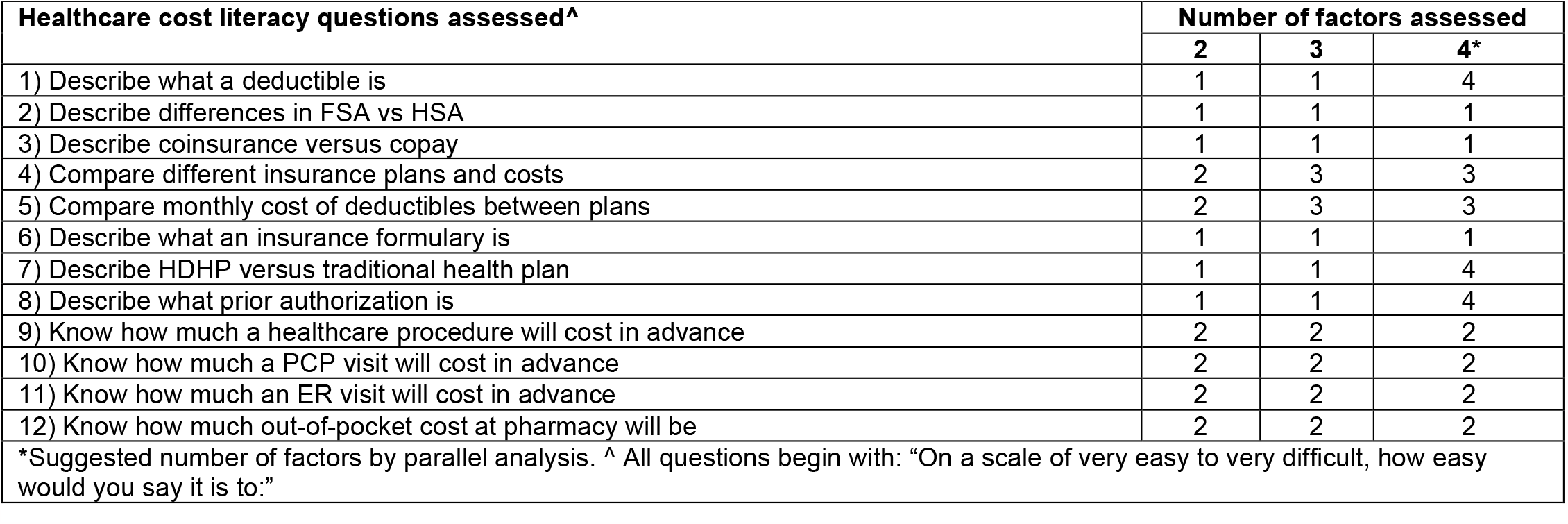
Factor assignment of healthcare cost literacy (HCL) questions.

**Table 2c.**
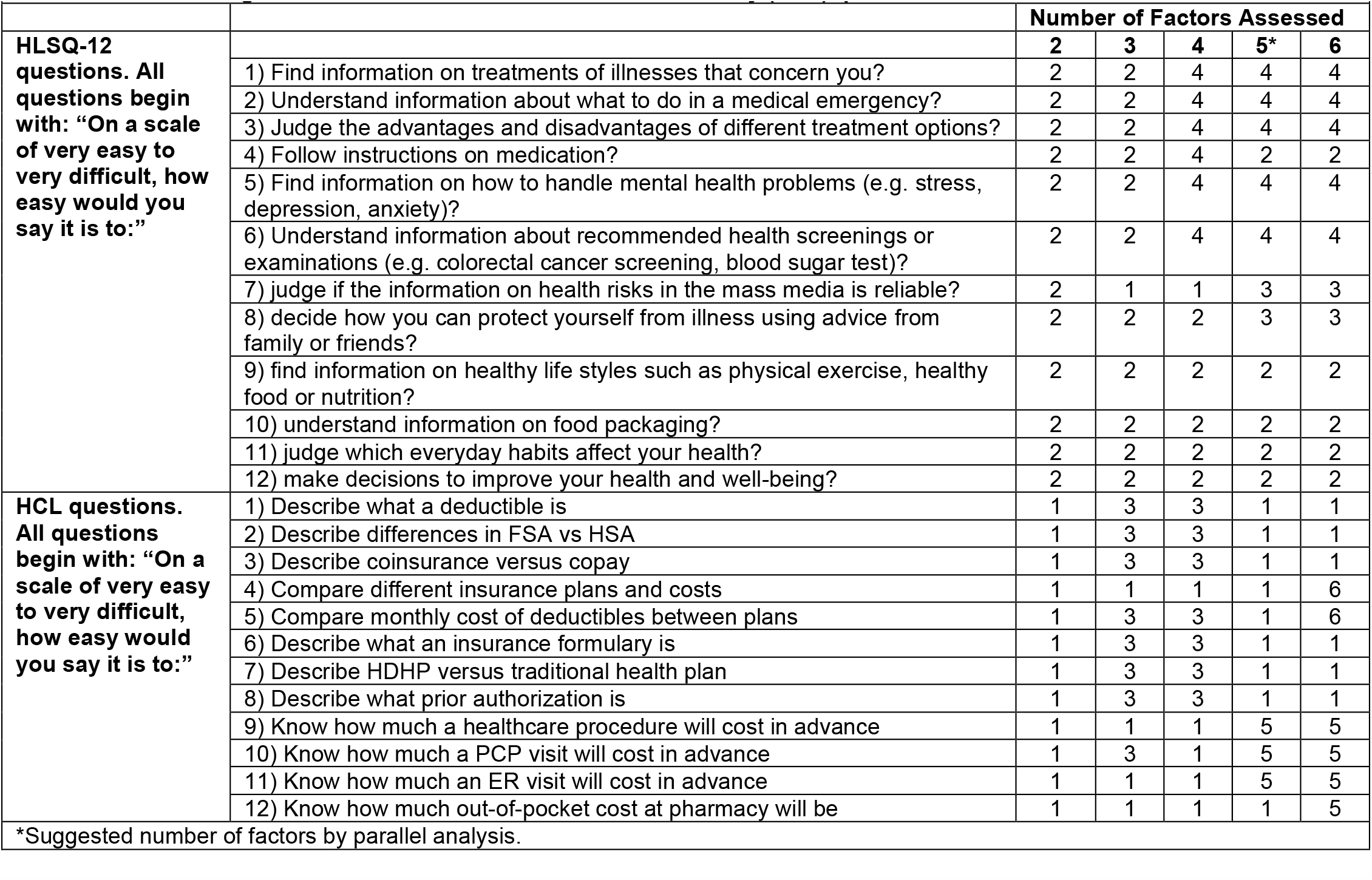
Factor assignment of HLSQ-12 and Healthcare cost literacy (HCL) questions.

The constructs that separated HCL into groups appear to be knowledge on health insurance terminology/interpretation (Factors 1 and 4), anticipatory expense assessment (Factor 2), and insurance plan comparison (Factor 3) **(Table 2b)**. Factor loadings ranged from 0.5 to 0.7, indicating strong fit on each of the three factors^15^. All three factors had sum of squared loadings greater than 1 (range: 1.69-2.30). When HL and HCL questions were grouped together **(Table 2c)**, the range of factor loadings was wider, from 0.4 to 0.7.

**Table 3a and 3b** describe mean HLSQ-12 and HCL questions across the sample accounting for imputed and non-imputed values.

**Table 3a.** Mean HL, Mean HCL across sample after removing individuals with >25% missing.

|  |  |  |  |  |  |
| --- | --- | --- | --- | --- | --- |
| HLSQ-12 N | Mean HL, Unweighted (SE) | Mean HL, Weighted (SE) | HCL N | Mean HCL, Unweighted (SE) | Mean HCL, Weighted (SE) |
| 1411 | 2.99 (0.01) | 2.98 (0.01) | 1298 | 2.44 (0.02) | 2.42 (0.02) |

**Table 3b.** Mean HL, Mean HCL across sample for individuals with responses (non-imputed) for all 24 HL+HCL questions.

|  |  |  |  |  |  |
| --- | --- | --- | --- | --- | --- |
| HLSQ-12 N | Mean HL, Unweighted (SE) | Mean HL, Weighted (SE) | HCL N | Mean HCL, Unweighted (SE) | Mean HCL, Weighted (SE) |
| 787 | 2.98 (0.02) | 2.97 (0.02) | 787 | 2.47 (0.02) | 2.44 (0.03) |

**Tables 4a and 4b** display results from a post-hoc sensitivity analysis on the EFA; mean scores across all three factors were higher among those who reported affirmatively to “Did you know that your hospital has a list of prices for services and items that is available for you to review in advance/front?”, “did you know that different pharmacies may charge different prices for the exact same prescription drug?”, and “did you know that it is possible to contest or negotiate a bill related to your healthcare (e.g. a doctor’s visit, a hospital procedure, etc.)?” versus those that answered “no”.

**Table 4a.** Association of binary questions on healthcare-related financial planning and 4 HCL factors (F1, F2, F3, F4)^ as informed by EFA.

|  |  |  |  |  |  |
| --- | --- | --- | --- | --- | --- |
|  |  | Mean F1* (SD) | Mean F2* (SD) | Mean F3* (SD) | Mean F4* (SD) |
|  | N | 1386 | 1451 | 1436 | 1464 |
| **Sample Mean** |  | 2.29 (0.81) | 2.40 (0.78) | 2.38 (0.81) | 2.72 (0.76) |
| **Did you know that your hospital has a list of prices for services and items that is available for you to review in advance/upfront?** |  |  |  |  |  |
| No | 1003 | 2.11 (0.77) | 2.24 (0.76) | 2.23 (0.78) | 2.58 (0.76) |
| Yes | 497 | 2.65 (0.78) | 2.70 (0.73) | 2.65 (0.80) | 2.98 (0.69) |
| **Did you know that different pharmacies may charge different prices for the exact same prescription drug?** |  |  |  |  |  |
| No | 328 | 2.05 (0.76) | 2.27 (0.76) | 2.23 (0.79) | 2.47 (0.75) |
| Yes | 1172 | 2.36 (0.82) | 2.43 (0.78) | 2.42 (0.82) | 2.79 (0.75) |
| **Did you know that it is possible to contest or negotiate a bill related to your healthcare (e.g. a doctor's visit, a hospital procedure, etc.)?** |  |  |  |  |  |
| No | 668 | 2.05 (0.76) | 2.21 (0.77) | 2.17 (0.79) | 2.50 (0.76) |
| Yes | 832 | 2.48 (0.80) | 2.55 (0.76) | 2.54 (0.80) | 2.89 (0.72) |
| ^ F1/F4 = Knowledge on health insurance terminology/interpretation<br>F2 = Anticipatory expense assessment<br>F3 = Insurance plan comparison<br>*A factor value is calculated for each observation where at least one of the factor components was answered other than 'Not sure'. Presented means are weighted. All p-values <.01 |  |  |  |  |  |

**Table 4b.** Association of binary questions on healthcare-related financial planning and 5 HL and HCL Factors (F1, F2, F3, F4, F5)^ as informed by EFA.

|  |  | Mean F1*<br>(SD) | Mean F2*<br>(SD) | Mean F3*<br>(SD) | Mean F4*<br>(SD) | Mean F5*<br>(SD) |
| --- | --- | --- | --- | --- | --- | --- |
|  | **N** | 1479 | 1487 | 1434 | 1471 | 1409 |
| **Sample Mean** |  | 2.49 (0.68) | 3.12 (0.52) | 2.64 (0.74) | 2.94 (0.59) | 2.32 (0.83) |
| **Did you know that your hospital has a list of prices for services and items that is available for you to review in advance/upfront?** |  |  |  |  |  |  |
| No | 1003 | 2.35 (0.66) | 3.08 (0.53) | 2.54 (0.74) | 2.87 (0.59) | 2.14 (0.81) |
| Yes | 497 | 2.78 (0.63) | 3.20 (0.51) | 2.82 (0.70) | 3.06 (0.55) | 2.65 (0.77) |
| **Did you know that different pharmacies may charge different prices for the exact same prescription drug?** |  |  |  |  |  |  |
| No | 328 | 2.29 (0.66) | 3.00 (0.53) | 2.52 (0.74) | 2.73 (0.61) | 2.19 (0.80) |
| Yes | 1172 | 2.55 (0.68) | 3.15 (0.52) | 2.67 (0.74) | 3.00 (0.57) | 2.35 (0.84) |
| **Did you know that it is possible to contest or negotiate a bill related to your healthcare (e.g. a doctor's visit, a hospital procedure, etc.)?** |  |  |  |  |  |  |
| No | 668 | 2.28 (0.66) | 3.02 (0.52) | 2.54 (0.74) | 2.79 (0.59) | 2.10 (0.80) |
| Yes | 832 | 2.66 (0.65) | 3.20 (0.51) | 2.71 (0.73) | 3.05 (0.56) | 2.49 (0.81) |

^ F1 = Knowledge on health insurance terminology/interpretation (HCL)
F2 = HLSQ-12 Questions 4, 9-12
F3 = HLSQ-12 Questions 7-8
F4 = HLSQ-12 Questions 1-3, 5-6
F5 = Anticipatory cost assessment (HCL)
\* A factor value is calculated for each observation where at least one of the factor components was answered other than 'Not sure'. Presented means are weighted. All p-values <.01

The results of the 5-item assessment on health insurance across the sample are included in **Table 5**. The percentage answering each question correctly ranged from 36% to 86%.

**Table 5.** Health insurance knowledge assessment and percentage correct across sample*.

| Question | Answer choices (*correct*) | Weighted percent correct |
| --- | --- | --- |
| If an insurance policy has a higher deductible, the premium should be lower, all else equal | *True*/False | 61.7 |
| If you visit a doctor who is not part of your insurer's network, you will generally have to pay more out of pocket | *True*/False | 86.4 |
| Generic prescription drugs generally cost the patient more than the brand name drugs | True/*False* | 79.9 |
| Which type of insurer places greater restrictions on patients' choices of the providers they see? | *HMO* / PPO / They are the same | 36.4 |
| Which of the following best describes a deductible? | A) A small amount that patients must pay each time they visit a doctor<br>B) *The amount patients must pay during a year before their insurance will pay for care*<br>C) The price policy holders must pay for insurance | 57.2 |

\*All assessment questions also included an option choice "not sure", coded as incorrect. Values are weighted.

The mean scores for the four-item assessment, HL, and HCL across various demographic variables and health management variables are presented in **Table 6**. Mean scores on the health insurance assessment, HL, and HCL were all higher with increasing age, better self-rated health status, and more confidence with healthcare management.

**Table 6.** Mean Assessment, Mean HL and Mean HCL across selected covariates.

| Variable |  | Mean Assessment Score (SE) | p | Mean HL Score (SE) | p | Mean HCL Score (SE) | p |
| --- | --- | --- | --- | --- | --- | --- | --- |
| Mean Assessment Score | < Median |  |  | 2.95 (0.02) | 0.03 | 2.36 (0.03) | <.01 |
|  | > Median |  |  | 3.01 (0.02) |  | 2.48 (0.03) |  |
| Mean HL Score | < Median | 3.22 (0.06) | 0.11 |  |  | 2.12 (0.02) | <.01 |
|  | > Median | 3.35 (0.05) |  |  |  | 2.78 (0.03) |  |
| Mean HCL Score | < Median | 3.22 (0.06) | <.01 | 2.72 (0.02) | <.01 |  |  |
|  | > Median | 3.44 (0.05) |  | 3.19 (0.02) |  |  |  |
| Age | 18-39 | 2.67 (0.07) | <.01 | 2.93 (0.03) | <.01 | 2.35 (0.04) | <.01 |
|  | 40-64 | 3.45 (0.06) |  | 2.99 (0.02) |  | 2.41 (0.03) |  |
|  | 65+ | 3.74 (0.06) |  | 3.05 (0.03) |  | 2.59 (0.04) |  |
| Gender | Female | 3.23 (0.05) | 0.67 | 3.01 (0.02) | 0.02 | 2.43 (0.03) | 0.91 |
|  | Male | 3.20 (0.06) |  | 2.95 (0.02) |  | 2.42 (0.03) |  |
| Race/Ethnicity | Black | 2.69 (0.12) | <.01 | 3.13 (0.04) | <.01 | 2.62 (0.06) | <.01 |
|  | Hispanic | 2.57 (0.12) |  | 2.92 (0.04) |  | 2.34 (0.07) |  |
|  | White | 3.49 (0.04) |  | 2.97 (0.02) |  | 2.41 (0.02) |  |
|  | Other | 3.16 (0.14) |  | 2.98 (0.05) |  | 2.38 (0.07) |  |
| Educational Attainment | HS or Less | 2.77 (0.07) | <.01 | 2.98 (0.02) | 0.77 | 2.40 (0.03) | 0.38 |
|  | More than HS | 3.49 (0.04) |  | 2.97 (0.02) |  | 2.44 (0.03) |  |
| Employment Status | Full-Time | 3.34 (0.07) | <.01 | 2.97 (0.02) | <.01 | 2.45 (0.03) | <.01 |
|  | Part-Time | 2.93 (0.09) |  | 2.89 (0.03) |  | 2.33 (0.07) |  |
|  | Retired | 3.75 (0.06) |  | 3.08 (0.03) |  | 2.61 (0.04) |  |
|  | Unemployed | 2.45 (0.18) |  | 2.89 (0.06) |  | 2.33 (0.09) |  |
|  | Other | 2.90 (0.09) |  | 3.00 (0.03) |  | 2.27 (0.04) |  |
| Household Income | <30k | 2.70 (0.08) | <.01 | 2.99 (0.03) | 0.26 | 2.34 (0.06) | <.01 |
|  | 30k-100k | 3.29 (0.06) |  | 2.96 (0.02) |  | 2.41 (0.03) |  |
|  | 100k+ | 3.62 (0.07) |  | 3.01 (0.03) |  | 2.54 (0.04) |  |
| Insurance Status | Employer-sponsored insurance | 3.53 (0.06) | <.01 | 2.98 (0.02) | 0.28 | 2.40 (0.04) | <.01 |
|  | Insurance from exchange | 3.21 (0.14) |  | 2.97 (0.05) |  | 2.49 (0.07) |  |
|  | Medicaid | 2.69 (0.10) |  | 2.95 (0.04) |  | 2.35 (0.06) |  |
|  | Medicare | 3.67 (0.06) |  | 3.03 (0.03) |  | 2.56 (0.04) |  |
|  | Other insurance | 3.19 (0.12) |  | 3.02 (0.05) |  | 2.36 (0.07) |  |
|  | Uninsured | 2.22 (0.11) |  | 2.92 (0.04) |  | 2.36 (0.05) |  |
| Self-rated Health | Good - Excellent | 3.25 (0.05) | 0.15 | 3.02 (0.02) | <.01 | 2.46 (0.02) | <.01 |
|  | Fair - Poor | 3.12 (0.08) |  | 2.86 (0.03) |  | 2.29 (0.04) |  |
| Confidence in ability to manage healthcare | Very confident-extremely confident | 3.39 (0.04) | <.01 | 3.10 (0.02) | <.01 | 2.65 (0.02) | <.01 |
|  | Not confident at all – little confident | 3.02 (0.07) |  | 2.67 (0.02) |  | 1.88 (0.03) |  |
| Confidence in ability to schedule healthcare appointments/follow-ups | Very comfortable - extremely comfortable | 3.40 (0.04) | <.01 | 3.10 (0.02) | <.01 | 2.65 (0.02) | <.01 |
|  | Not comfortable at all – little comfortable | 3.03 (0.07) |  | 2.72 (0.02) |  | 1.94 (0.03) |  |
| Know at what age to get screened for cancer | Yes | 3.43 (0.05) | <.01 | 3.05 (0.02) | <.01 | 2.55 (0.02) | <.01 |
|  | No | 2.87 (0.08) |  | 2.82 (0.03) |  | 2.13 (0.04) |  |
| Completed a wellness visit in last 12mos | Yes | 3.36 (0.04) | <.01 | 3.00 (0.02) | 0.09 | 2.46 (0.02) | <.01 |
|  | No | 2.92 (0.09) |  | 2.94 (0.03) |  | 2.28 (0.04) |  |
| Compare insurance plans annually | Yes | 3.49 (0.05) | <.01 | 3.04 (0.02) | <.01 | 2.59 (0.02) | <.01 |
|  | No | 3.13 (0.06) |  | 2.93 (0.03) |  | 2.25 (0.03) |  |
| Received at least one COVID-19 vaccine | Yes | 3.31 (0.05) | <.01 | 2.99 (0.02) | 0.23 | 2.46 (0.02) | <.01 |
|  | No | 3.04 (0.08) |  | 2.95 (0.03) |  | 2.32 (0.04) |  |

## Discussion

Limited research has been done to assess current population-level understanding of the costs associated with medical care. In this study, we introduce HCL as a potential new mechanism for assessing individuals’ literacy on the costs associated with healthcare. We also assess how HL is associated with HCL and make distinctions between the two related, but unique, concepts. An understanding of Americans’ levels of HCL will help policymakers and actors in the healthcare system develop targeted plans to educate consumers on financial planning and evaluation in the healthcare system.

An EFA revealed four distinct underlying variables making up the HCL components we assessed. This potentially suggests several themes that researchers can use in future work in the areas of health insurance terminology awareness, healthcare cost planning, and insurance selection/planning. When HL and HCL Likert questions were grouped together, 5-factors were suggested to be optimal; conceptually, the 4-factor scale may be more consistent with the question groupings. Further research in this field should explore factor determinations on larger and/or more diverse samples to validate or provide additional guidance on these findings.

The four-item “assessment” given to participants centered on the HCL-related concept of health insurance revealed distinct differences across certain groups. People who had more interactions with the healthcare system (through receiving COVID-19 vaccination, and completing a wellness visit) had higher mean HCL scores on this assessment. This is consistent with historical literature showing that education on health insurance could be a useful tool in improving individuals’ understanding of their plans and healthcare options^16^. Further research should evaluate the relationship between HCL and ability to navigate and understand health insurance.

There are limitations worth noting in our approach. Firstly, the survey data is cross-sectional, and therefore we were unable to confirm causal relationships between health literacy and the medical cost literacy questions assessed. And as with any survey, there may be response biases present. This survey was only offered to English-speaking respondents, effectively excluding a portion of the US population. Despite its limitations, there are important strengths to this work. Importantly, this work is novel; research into cost literacy specifically associated with healthcare is very limited in nature. Secondly, the study is nationally representative allowing us to draw conclusions and generate hypotheses that are generalizable to the U.S.

This study is the first one to survey US adults on their familiarity and confidence with navigating the financial components of the healthcare system. Developing a further-validated measure of HCL for further research would help public health practitioners develop and evaluate interventions to improve HCL, especially for those who need it most.

The US healthcare system is convoluted and challenging to navigate; it is unsurprising that many individuals struggle with understanding the costs associated with medical care. Although research has shown that patients want to be informed about healthcare costs^17,18^, it is evident that there is work to be done in terms of increasing literacy around costs associated with healthcare – particularly for adults in middle-age and those with lower levels of health literacy.

## Data Availability

All data produced in the present work are contained in the manuscript.

## Notes

### Competing Interest Statement

The authors have declared no competing interest.

### Funding Statement

This study did not receive any external funding.

### Author Declarations

This study was determined exempt from IRB oversight by the Advarra IRB. Authors were provided access to de-identified survey responses by YouGov, who was responsible for collecting, storing and managing the survey responses. YouGov works with the Western IRB to ensure its research protocol and specific studies are consistent with Good Clinical Practices as defined under the U.S. Food and Drug Administration (FDA) regulations and the International Conference on Harmonisation (ICH) guidelines.

## References

1. Berkman ND, Sheridan SL, Donahue KE, Halpern DJ, Crotty K. Low Health Literacy and Health Outcomes: An Updated Systematic Review. Ann Intern Med. 2011;155(2):97. doi:10.7326/0003-4819-155-2-201107190-00005

2. Goggins K, Wallston K, Nwosu S, Schildcrout J, Castel L, Kripalani S. Health literacy, numeracy, and other characteristics associated with hospitalized patients’ preferences for involvement in decision-making. J Health Commun. 2014;19(0 2):29–43. doi:10.1080/10810730.2014.938841

3. Baker DW, Gazmararian JA, Williams MV, et al. Functional health literacy and the risk of hospital admission among Medicare managed care enrollees. Am J Public Health. 2002;92(8):1278–1283.

4. Levy H, Janke A. Health Literacy and Access to Care. J Health Commun. 2016;21(Suppl):43–50. doi:10.1080/10810730.2015.1131776

5. Sudore RL, Mehta KM, Simonsick EM, et al. Limited Literacy in Older People and Disparities in Health and Healthcare Access: limited literacy in older people and health disparities. J Am Geriatr Soc. 2006;54(5):770–776. doi:10.1111/j.1532-5415.2006.00691.x

6. Nutbeam D, McGill B, Premkumar P. Improving health literacy in community populations: a review of progress. Health Promot Int. 2018;33(5):901–911. doi:10.1093/heapro/dax015

7. Kountz DS. Strategies for Improving Low Health Literacy. Postgrad Med. 2009;121(5):171–177. doi:10.3810/pgm.2009.09.2065

8. Walters R, Leslie SJ, Polson R, Cusack T, Gorely T. Establishing the efficacy of interventions to improve health literacy and health behaviours: a systematic review. BMC Public Health. 2020;20(1):1040. doi:10.1186/s12889-020-08991-0

9. Cunningham PJ. The Growing Financial Burden Of Health Care: National And State Trends, 2001–2006. Health Aff (Millwood). 2010;29(5):1037–1044. doi:10.1377/hlthaff.2009.0493

10. Banthin J, Cunningham P, Bernard D. Financial Burden Of Health Care, 2001 2004. HealthAff Proj Hope. 2008;27:188–195. doi:10.1377/hlthaff.27.1.188

11. Paez K, Zhao L, Hwang W. Rising Out-Of-Pocket Spending For Chronic Conditions: A Ten-Year Trend. Health Aff (Millwood). 2009;38:15–25. doi:10.1377/hlthaff.28.1.15

12. Finbråten HS, Wilde-Larsson B, Nordström G, Pettersen KS, Trollvik A, Guttersrud Ø. Establishing the HLS-Q12 short version of the European Health Literacy Survey Questionnaire: latent trait analyses applying Rasch modelling and confirmatory factor analysis. BMC Health Serv Res. 2018;18:506. doi:10.1186/s12913-018-3275-7

13. Understanding America Study. https://uasdata.usc.edu/index.php

14. Çokluk, Omay, Koçak, Duygu. Using Horn’s Parallel Analysis Method in Exploratory Factor Analysis for Determining the Number of Factors. Educ Sci Theory Pract. Published online 2016. doi:10.12738/estp.2016.2.0328

15. Howard, Matt C. A Review of Exploratory Factor Analysis Decisions and Overview of Current Practices: What We Are Doing and How Can We Improve? 10.1080/10447318.2015.1087664

16. Marquis MS. Consumers’ Knowledge about their Health Insurance Coverage. Health Care Financ Rev. 1983;5(1):65–80.

17. Alexander GC, Casalino LP, Meltzer DO. Patient-Physician Communication About Out-of-Pocket Costs. JAMA. 2003;290(7):953–958. doi:10.1001/jama.290.7.953

18. Ahmed IA, Harvey A, Amsellem M, Smith TJ. Provider-patient communication about cost of care: Results from a national patient education program. J Clin Oncol. 2013;31(15_suppl):9578–9578. doi:10.1200/jco.2013.31.15_suppl.9578

